# Predictive Analysis of COVID-19 Spread in Sri Lanka using an Adaptive Compartmental Model: Susceptible-Exposed-Infected-Recovered-Dead (SEIRD) Model

**DOI:** 10.1101/2021.08.09.21261819

**Authors:** R. M. Nayani Umesha Rajapaksha, Millawage Supun Dilara Wijesinghe, Toms K. Thomas, Sujith P. Jayasooriya, B. M. W. Indika Gunawardana, W. M. Prasad Chathuranga Weerasinghe, Shalini Bhakta, Yibeltal Assefa

## Abstract

Novel Corona Virus (COVID-19) is still spreading throughout the world despite various degrees of movement restrictions and the availability of multiple safe and effective vaccines. Modelling in predicting the spread of an epidemic is important for health planning and policies. This study aimed to apply a dynamic Susceptible-Exposed-Infected-Recovered-Deaths (SEIRD) model and simulated it under a range of epidemic conditions using python programme language. The predictions were based on different scenarios from without any preventive measures to several different preventive measures under R_0_ of 4. The model shows that more weight to personal protection can halt the spread of transmission followed by the closure of public places and interprovincial movement restriction. Results after simulating various scenarios indicate that disregarding personal protective measures can have devastating effects on the local population. Strict adherence, maintaining and monitoring of self-preventive measures are vital towards minimizing the death toll from COVID-19.

## Introduction

The coronavirus disease has become a pandemic that poses a serious public health risk globally. The virus is mutating rapidly and producing many strains, which are a significant threat to the control measures. The alpha strain is a more transmissible variant initially detected in the United Kingdom (UK) and circulating in Sri Lanka as the main variant (Economy next, 2021). However, the delta variant which has higher transmissibility is being detected in several places. In this context, an in-depth understanding of the current epidemic and demand dynamics is fundamental in health planning and policymaking, especially when the resources are limited. With the purpose of forecasting, different prediction models are proposed by various academics and groups (Kumar, 2017; Sperrin & McMillan, 2020). Compartmental models can be used to project scenarios with various disease control measures individually or as a useful combination for evidence-based policy formulation. Furthermore, epidemiologists have been using mass action, compartmental models, over a hundred years which are famous for simplicity in both analysis and outcome assessment (Adiga et al., 2020). One scientific way of predicting the future directions and trends of an epidemic is the development of different compartment models (He et al., 2020). The key element in this field of research is being able to link mathematical models and data. Both epidemiological data and findings of mathematical model studies can be compared for optimal results and guidance. Mathematical modelling plays a vital role in the highest level of policymaking in the fields of health economics, emergency planning, monitoring of surveillance data and risk assessment and control. This paper aims to describe a dynamic Susceptible-Exposed-Infected-Recovered-Deaths (SEIRD) model and simulate it under a range of epidemiological conditions to give an insight into COVID-19 spread in Sri Lanka.

## Methods

Many compartmental models belong to the basic Susceptible-Infectious-Recovered (SIR) class (Adiga et al., 2020; El-Doma, 2006; Youssef et al., 2020). The SIR models are further extended by adding an Exposed (E) compartment. We constructed a compartmental epidemiological model in the Figure 1 with vital dynamics describing the number of individuals in a fixed population who are susceptible to infection (S), exposed (E), infected (I), recovered (R), and deaths (D) compartments(Adiga et al., 2020; He et al., 2020).

**Figure 1.**
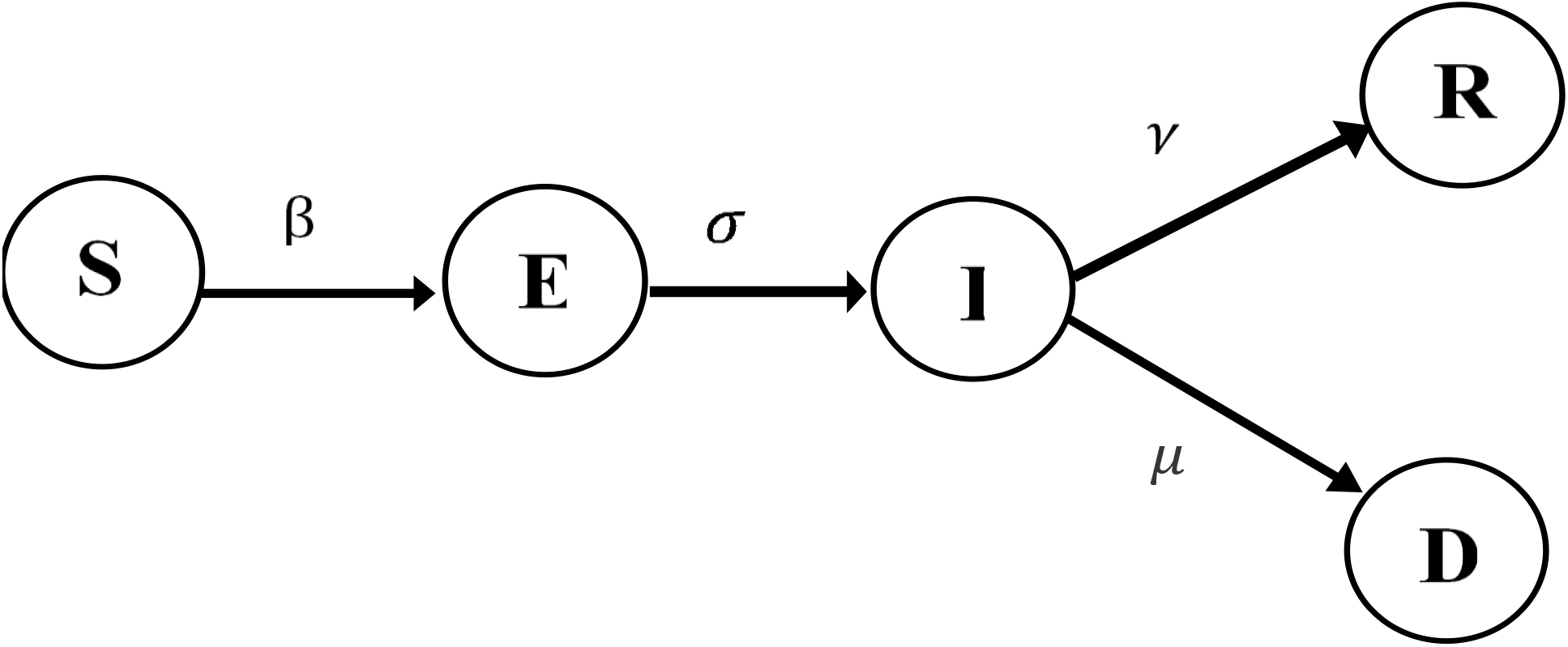
SEIRD Model with Transition Forces.

**Figure 2.**
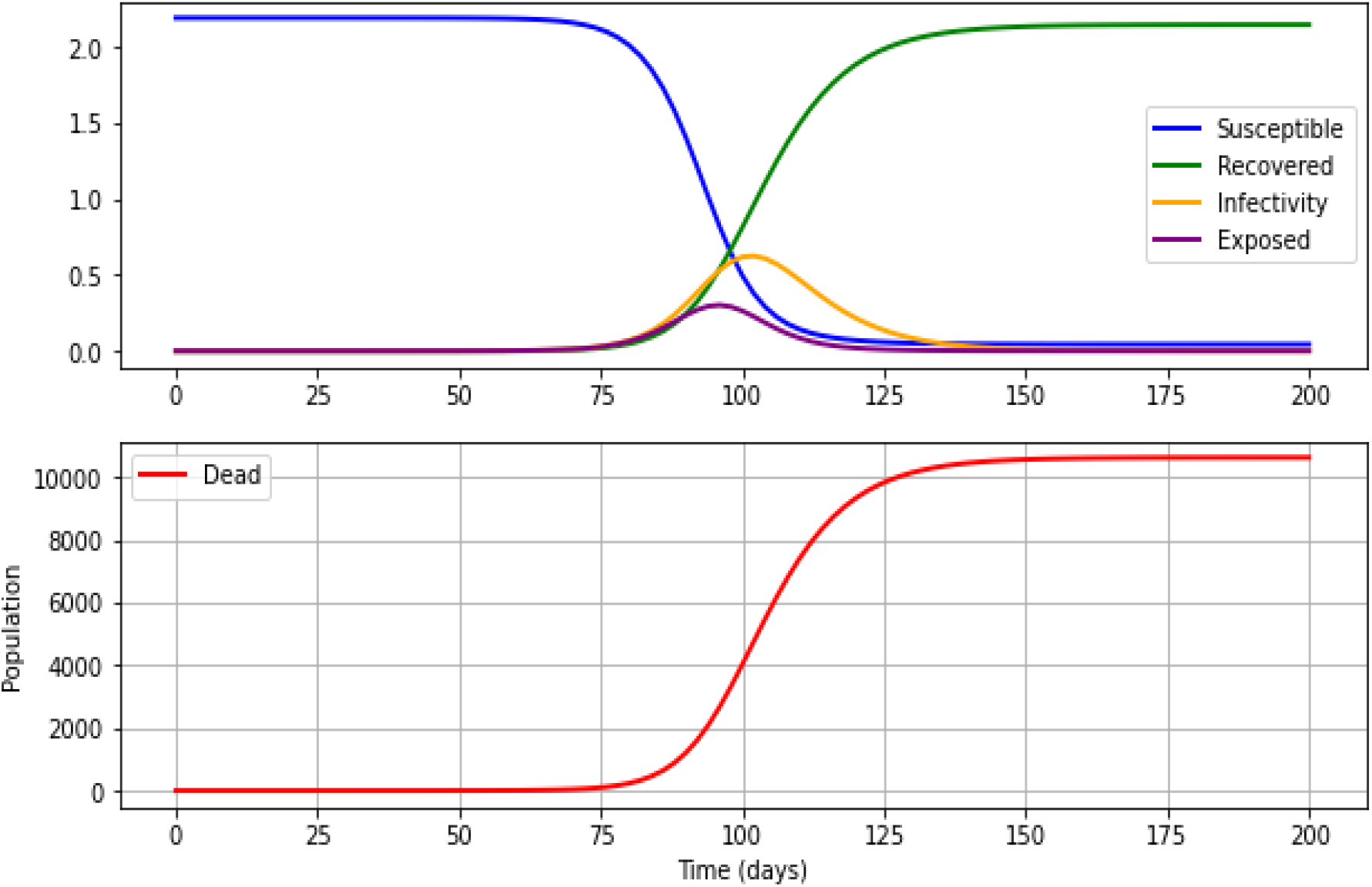
Predictions based on the SEIRD model R_0_ = 4 without any preventive strategies. X axis: Population= e10^7^

We extracted publicly available data with permission from the official website of the Health Promotion Bureau (HPB) and the Epidemiology Unit, Ministry of Health, Sri Lanka (Epidemiology Unit - Ministry of Health Sri Lanka, 2021; Health Promotion Bureau - Ministry of Health, 2021). We used anonymized data for this analysis and extracted data relevant to cases reported from the 11^th^ of March 2020 to the 5^th^ of July 2021. For the development of the prediction model, three dynamic variables were considered. The first variable was personal measures (the practice of social distancing, wearing masks and handwashing) which was considered by the way they were adopted (100%, 50% and 25%). The second variable was inter-provincial movement restrictions with 100%, 75% and 50% adaptation. The third variable was the closure of places (public place, school, workplace) with 100%, 50% and 33% adaptation. The weighted factors for the three scenarios were 0.70, 0.33, 0.10 for personal measures, 0.15, 0.33, 0.60 for the movement restrictions, and 0.15, 0.33 and 0.30 for the closure of places, respectively. The predictions for SEIRD were made when the R_0_ value is 4. Python programming language was used for the analysis.

### Model equations

The flow of individuals through the compartments of the model is governed by a set of Ordinary Differential Equations (ODE) as given below.

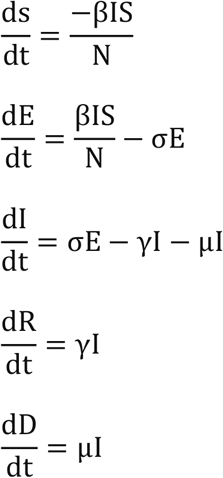

### Disease characteristics and model parameters

The available COVID-19 data was used as the disease characteristics in this exploration and the model parameters were based on the data (Department of Census and Statistics - Sri Lanka, 2021; Epidemiology Unit - Ministry of Health Sri Lanka, 2021; He et al., 2020; Moss et al., 2020; Verity et al., 2020; World Health Organization, 2020a, 2020b, 2021).

## Results

The figures below show the predictions of various preventive strategies which were based on R_0_ of 4. The X-axis shows the time (in days), and the Y-axis shows the population. The number of susceptible individuals is shown in blue, recovered in green, infective in yellow, exposed in purple and deaths in red.

### Predictions based on the SEIRD model without any preventive strategies

If the R_0_=4, the peak of the infectious will occurs around day 100 with 6.3 million infected individuals. Out of all exposed and infected individuals, 10,617 will die following 150 days after the beginning of the epidemic curve if there are no proper strategies to prevent the outbreak.

**Predictions of SEIRD with 70% of personal protection (social distancing, wearing masks and handwashing), 15% of movement restrictions and 15% of closure places (public place, school, workplace) with R**_**0**_ **of 4 with different strategies**

We observed how the SEIRD dynamics are affected by different preventive strategies for COVID-19 at a specific time in the system’s evolution.

### With the implementation of 100%, 50% and 25% of personal protection

With 100%, 50% and 25% of personal protection are implemented at R_0_ of 4, the rate of deaths will be increased from 600 days, 125 days, 85 days, with 3,409 deaths in 1200 days (with 0.24 million infected individuals), 9,803 deaths in 250 days (with 3.9 million infected individuals) and 10,373 deaths in 175 days (with 5.2 million infected individuals) will be observed, respectively. There is no visible peak observed with 100% personal protection. However, the number of days to achieve the peak of the infection curve will be 160 days with 3 million and 125 days with 4 million infected individuals at the peak of the infection curves with 50% and 25% personal protection, respectively. Additional file shows in more details. Please refer figure 3 given below and table 2.

**Table 1.**
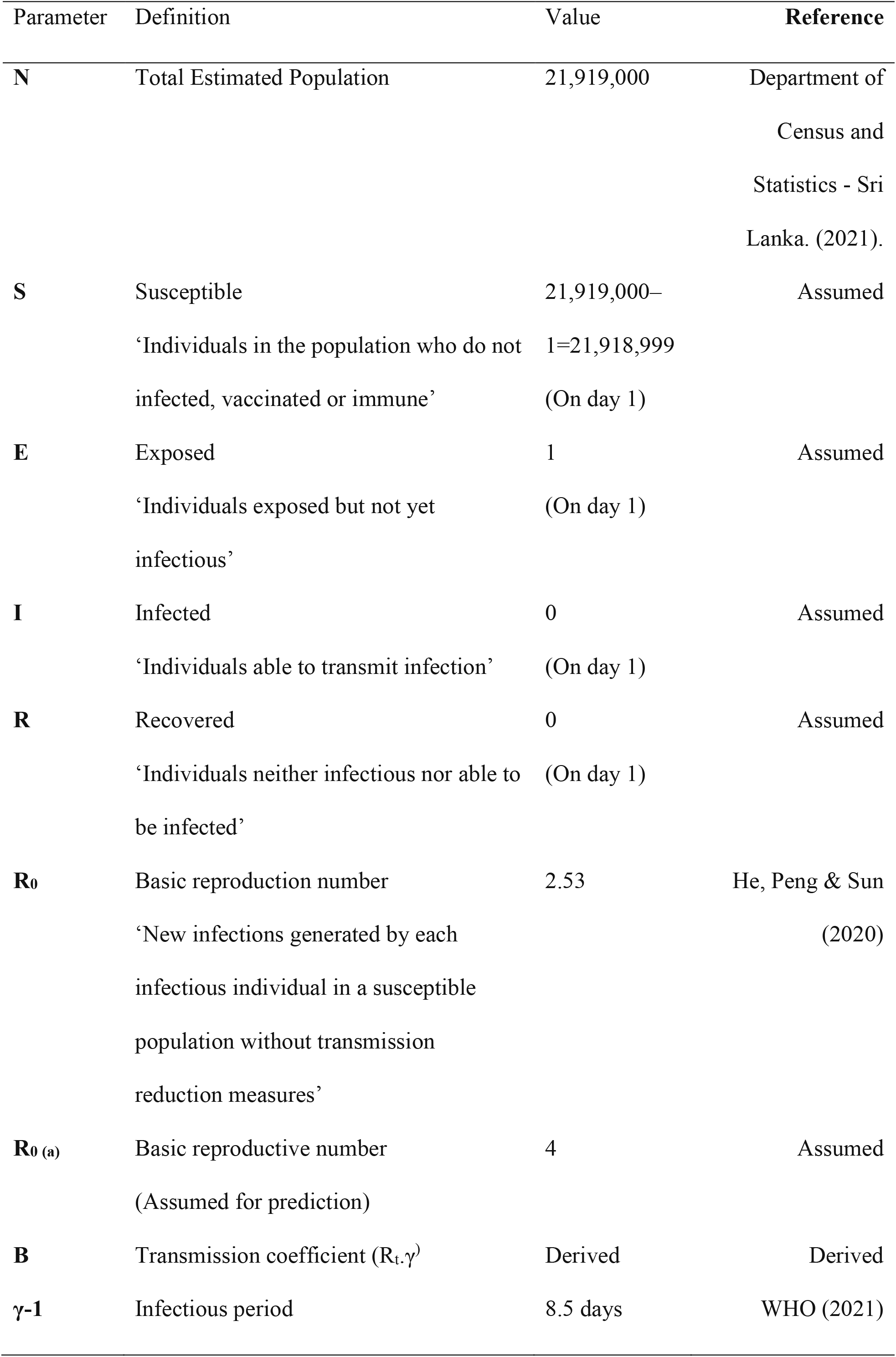
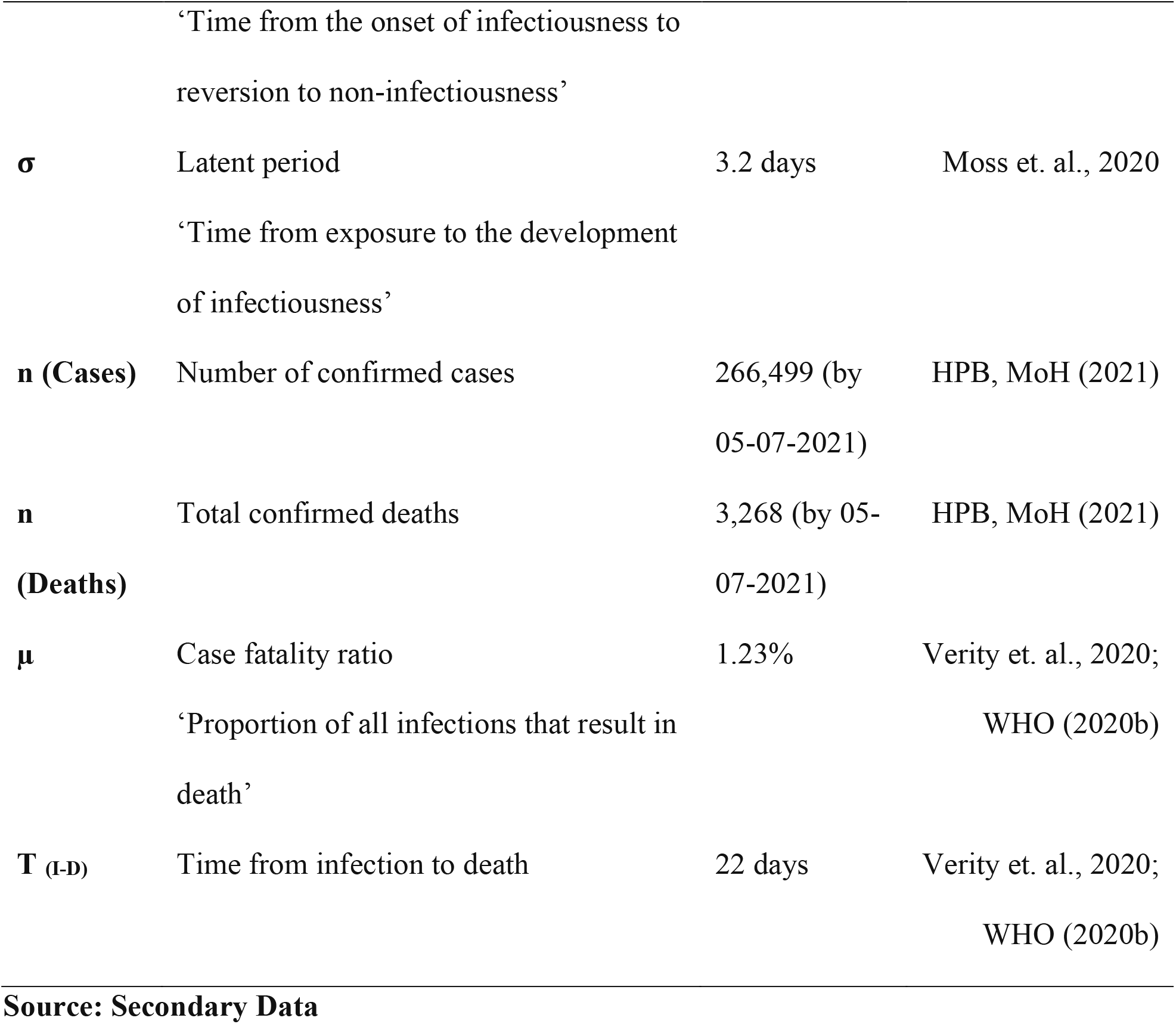
Disease characteristics and model parameters.

| Parameter | Definition | Value | Reference |
| --- | --- | --- | --- |
| **N** | Total Estimated Population | 21,919,000 | Department of<br>Census and<br>Statistics - Sri<br>Lanka. (2021). |
| **S** | Susceptible<br>‘Individuals in the population who do not<br>infected, vaccinated or immune’ | 21,919,000–<br>1=21,918,999<br>(On day 1) | Assumed |
| **E** | Exposed<br>‘Individuals exposed but not yet<br>infectious’ | 1<br>(On day 1) | Assumed |
| **I** | Infected<br>‘Individuals able to transmit infection’ | 0<br>(On day 1) | Assumed |
| **R** | Recovered<br>‘Individuals neither infectious nor able to<br>be infected’ | 0<br>(On day 1) | Assumed |
| **R<sub>0</sub>** | Basic reproduction number<br>‘New infections generated by each<br>infectious individual in a susceptible<br>population without transmission<br>reduction measures’ | 2.53 | He, Peng & Sun<br>(2020) |
| **R<sub>0</sub> (a)** | Basic reproductive number<br>(Assumed for prediction) | 4 | Assumed |
| **B** | Transmission coefficient ($R_t \cdot \gamma$) | Derived | Derived |
| **$\gamma^{-1}$** | Infectious period | 8.5 days | WHO (2021) |
|  | ‘Time from the onset of infectiousness to<br>reversion to non-infectiousness’ |  |  |
| $\sigma$ | Latent period | 3.2 days | Moss et. al., 2020 |
|  | ‘Time from exposure to the development<br>of infectiousness’ |  |  |
| **n (Cases)** | Number of confirmed cases | 266,499 (by<br>05-07-2021) | HPB, MoH (2021) |
| **n (Deaths)** | Total confirmed deaths | 3,268 (by 05-07-2021) | HPB, MoH (2021) |
| $\mu$ | Case fatality ratio | 1.23% | Verity et. al., 2020; |
|  | ‘Proportion of all infections that result in<br>death’ |  | WHO (2020b) |
| **T (I-D)** | Time from infection to death | 22 days | Verity et. al., 2020; |
|  |  |  | WHO (2020b) |

**Source: Secondary Data**

**Table 2.** Predictions of SEIRD: The following table give a summary of SEIRD model prediction without any measures and with all three strategies [personal protection, movement restrictions and closure of places].

| Options | Weight | Policy | Death | Days | Exposed | Infected |
| --- | --- | --- | --- | --- | --- | --- |
| No measures | - | Without | 10617 | 150 | 3020859 | 6252900 |
| **SCENARIO 01** |  |  |  |  |  |  |
| Options | Weight | Policy | Death | Days | Exposed | Infected |
| Personal Measures | 0.7 | 100 | 3409 | 1200 | 89317 | 236313 |
|  |  | 50 | 9803 | 250 | 1670828 | 3903499 |
|  |  | 25 | 10373 | 175 | 2386508 | 5230010 |
| Movement Restrictions | 0.15 | 100 | 10417 | 175 | 2474203 | 5384452 |
|  |  | 75 | 10481 | 160 | 2625315 | 5635264 |
|  |  | 50 | 10535 | 150 | 2755947 | 5845803 |
| Closure | 0.15 | 100 | 10417 | 175 | 2474203 | 5384452 |
|  |  | 50 | 10535 | 150 | 2755947 | 5845803 |
|  |  | 33 | 10566 | 145 | 2841936 | 5989039 |

| SCENARIO 02 |  |  |  |  |  |  |
| --- | --- | --- | --- | --- | --- | --- |
| Options | Weight | Policy | Death | Days | Exposed | Infected |
| Personal Measures | 0.33 | 100 | 9897 | 200 | 1758096 | 4072475 |
|  |  | 50 | 10387 | 175 | 2417448 | 5295067 |
|  |  | 25 | 10525 | 150 | 2732444 | 5810125 |
| Movement Restrictions | 0.33 | 100 | 9897 | 200 | 1758096 | 4072475 |
|  |  | 75 | 10165 | 200 | 2095694 | 4727986 |
|  |  | 50 | 10387 | 175 | 2417448 | 5295067 |
| Closure | 0.33 | 100 | 9897 | 200 | 1758096 | 4072475 |
|  |  | 50 | 10387 | 175 | 2417448 | 5295067 |
|  |  | 33 | 10487 | 150 | 2634210 | 5651165 |
| SCENARIO 03 |  |  |  |  |  |  |
| Options | Weights | Policy | Death | Days | Exposed | Infected |
| Personal Measures | 0.1 | 100 | 10500 | 150 | 2668474 | 5706441 |
|  |  | 50 | 10565 | 150 | 2838678 | 5984604 |
|  |  | 25 | 10592 | 150 | 2935906 | 6129321 |
| Movement Restrictions | 0.6 | 100 | 6960 | 425 | 502580 | 1293874 |
|  |  | 75 | 9142 | 250 | 1216562 | 2960599 |
|  |  | 50 | 10020 | 200 | 1885260 | 4315111 |
| Closure | 0.3 | 100 | 10020 | 200 | 1885260 | 4315111 |
|  |  | 50 | 10421 | 150 | 2474203 | 5384452 |
|  |  | 33 | 10503 | 150 | 2673467 | 5714203 |

**Source: Secondary Data from the Ministry of Health Sri Lanka and World Health Organization.**

**Figure 3.**
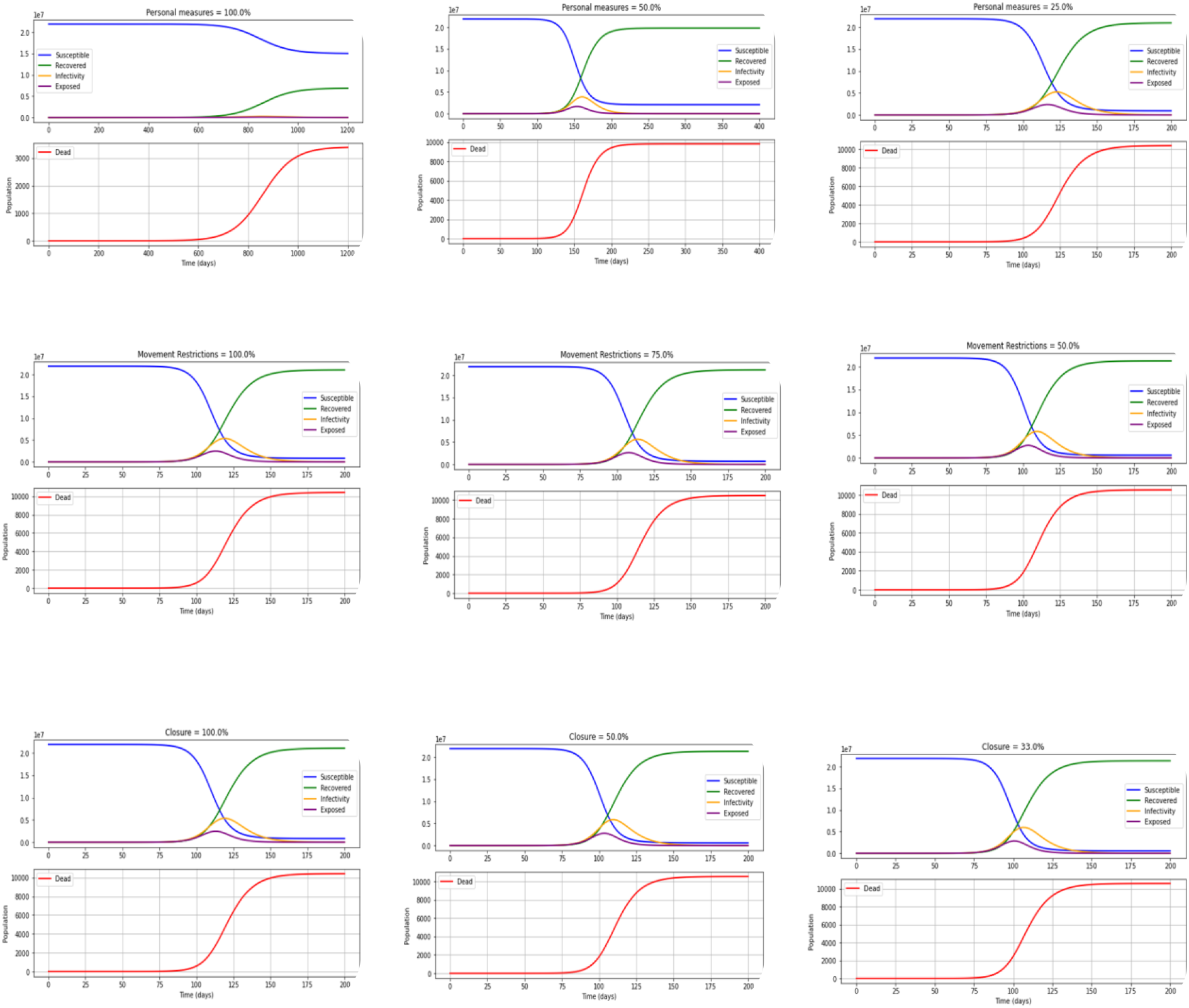
Predictions of SEIRD. The Figure below show the SEIRD predictions with 70% of personal protection, 15% of movement restrictions and 15% of closure places with Ro of 4 with different strategies

### With implementation of 100%, 75% and 50% of movement restrictions

With 100%, 75% and 50% of movement restrictions, the rate of deaths will be increased from 85 days, 80 days, and 75 days with 10,417 deaths in 175 days (with 5.4 million infected individuals), 10,481 deaths in 160 days (with 5.6 million infected individuals) and 10,535 deaths in 150 days (with 5.8 million infected individuals) will be observed, respectively. Moreover, the number of days to achieve the peak of the infection curve will be 120 days, 115 days, and 110 days, respectively. For more details refer figure 3 given above. Kindly refer the table below also for more detailed information on the various predictions.

### With the implementation of 100%, 50% and 33% of closure of places

With the 100%, 50% and 33% of closure of places are implemented at R_0_ of 4, the rate of deaths will be increased from 85 days, 80 days, and 75 days with 10,417 deaths in 175 days with 5.4 million infected individuals, 10,535 deaths in 150 days with 5.8 million infected individuals and 10,566 deaths in 145 days with 5.98 million infected individuals will be observed, respectively. Furthermore, the number of days to achieve the peak of the infection curve will be 120 days, 110 days, and 105 days with the infected individuals of approximately 4 million, 5 million and 6 million at the peak of the infection curves. See the figure 3 above for more details.

**Predictions of SEIRD with personal protection (social distancing, mask, hand washing), movement restrictions and closure (public place, school, workplace) with R**_**0**_ **of 4 with equal weight (33%) for all three dynamic variables**

### With the implementation of 100%, 50% and 25% of personal protection

With 100%, 50% and 25% of personal protection are implemented at R_0_ of 4, the rate of deaths will be increased from 120 days, 85 days, and 75 days and 9,897 deaths in 200 days (with 4.1 million infected individuals), 10,387 deaths in 175 days (with 5.3 million infected individuals) and 10,525 deaths in 150 days (with 5.8 million infected individuals) will be observed. Furthermore, the number of days to achieve the peak of the infection curve will be 150 days, 120 days, and 110 days with 4 million, 3.5 million and 5.8 million infected individuals at the peak of the infection curves respectively. The figure 3 and table 2 give more details

### With implementation of 100%, 75% and 50% of movement restrictions

With 100%, 75% and 50% of movement restrictions are implemented at R_0_ of 4, the rate of deaths will be increased from 115 days, 100 days, and 85 days with 9,663 deaths in 200 days (with 4.1 million infected individuals), 10,165 deaths in 200 days (with 4.7 million infected individuals) and 10,387 deaths in 175 days (with 5.3 million infected individuals) will be observed, respectively. Moreover, the number of days to achieve the peak of the infection curve will be 120 days, 115 days, and 110 days with an infected individuals of approximately 4 million, 4.5 million and 5 million at the peak of the infection curves, respectively. Refer figure 2 for more details along with the table 2.

### With the implementation of 100%, 50% and 33% of Closure of Places

With the 100%, 50% and 33% of closure of places are implemented at R_0_ of 4, the rate of deaths will be increased from 120 days, 90 days, and 80 days with 9,663 deaths in 200 days (with 4.1 million infected individuals), 10,387 deaths in 175 days (with 5.3 million infected individuals) and 10,487 deaths in 150 days (with 5.7 million infected individuals) will be observed. Furthermore, the number of days to achieve the peak of the infection curve will be 160 days, 120 days, and 115 days with an infected individuals of 2.5 million, 5 million and 5.5 million at the peak of the infection curves, respectively. Additional file shows in more details refer the Figure 4 table 2.

**Figure 4.**
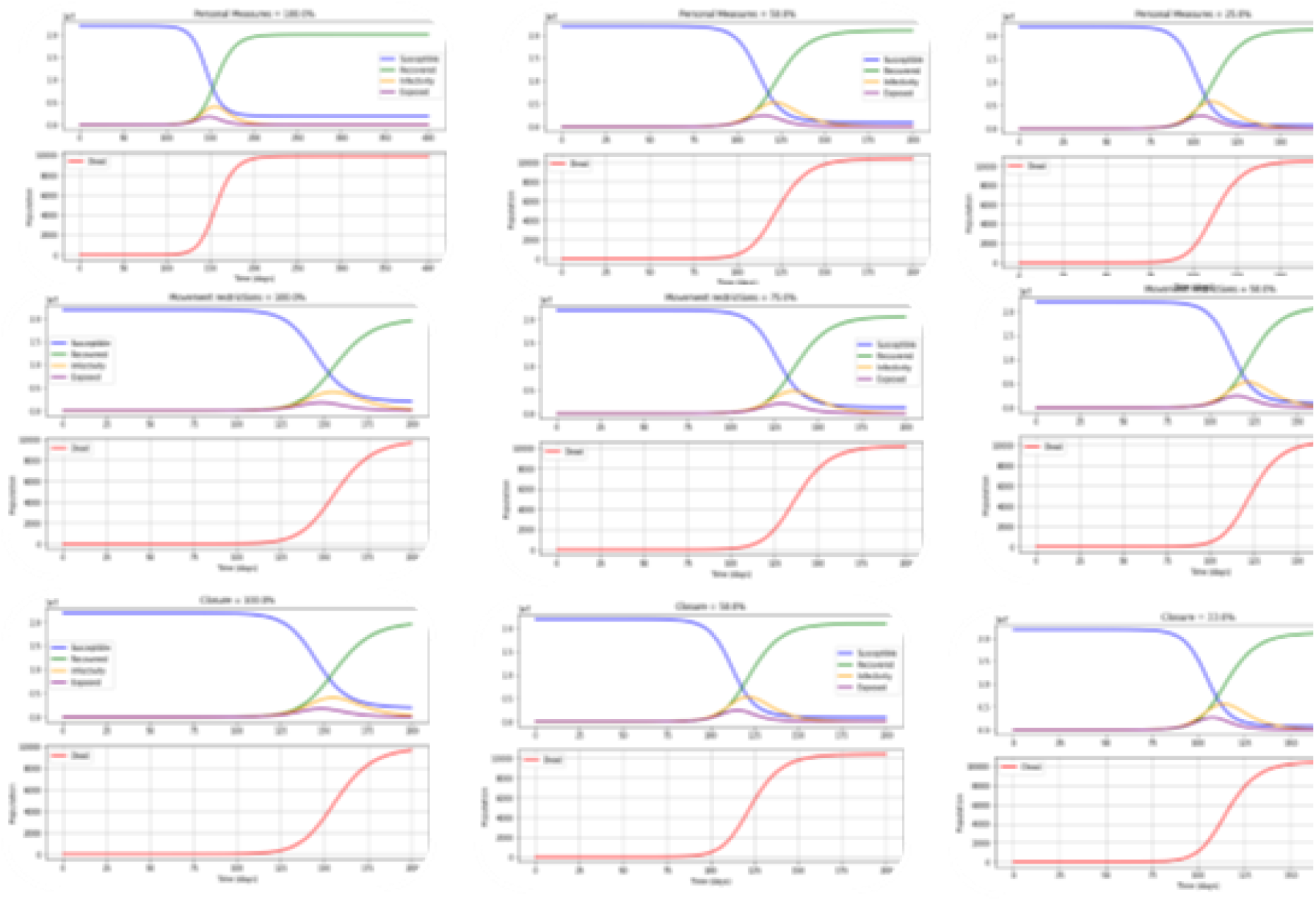
Predictions of SEIRD. The figure below shows the SEIRD prediction with personal protection, movement restrictions and closure of places with R_0_ of 4 with equal weight (33%) for all three dynamic variables

**Predictions of SEIRD with 10% of personal protection (social distancing, mask, hand washing), 60% of movement restrictions and 30% of closure of places (public place, school, workplace) with R**_**0**_ **of 4**

### With the implementation of 100%, 50% and 25% of personal protection

With the 100%, 50% and 25% of personal protection are implemented at R_0_ of 4, the rate of deaths will be increased from 85 days, 80 days, and 75 days, with 10,500 deaths in 150 days (with 6.3 million infected individuals), 10,565 deaths in 150 days (with 5.7 million infected individuals) and 10,592 deaths in 150 days (with 6.1 million infected individuals) will be observed, respectively. Furthermore, the number of days to achieve the peak of the infection curve will be 115 days, 110 days, and 105 days with 5 million, 6 million and 6.5 million infected individuals at the peak of the infection curves, respectively. Additional file shows in more details see figure 5 below and table 2.

**Figure 5.**
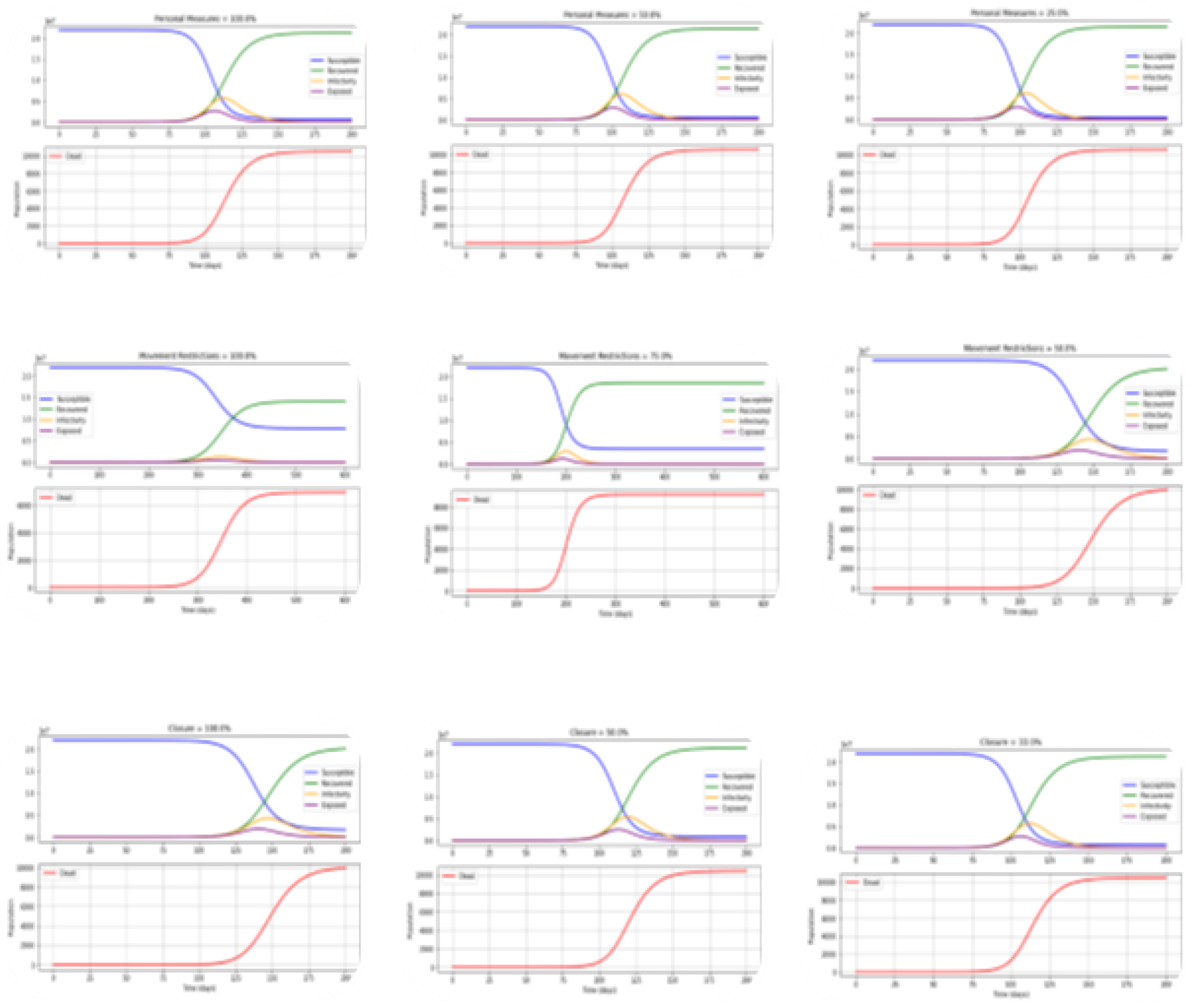
Predictions of SEIRD. The following table suggest the SEIRD prediction with 10% of personal protection, 60% of movement restrictions and 30% of closure of places with R_0_ of 4.

**Figure 6.**
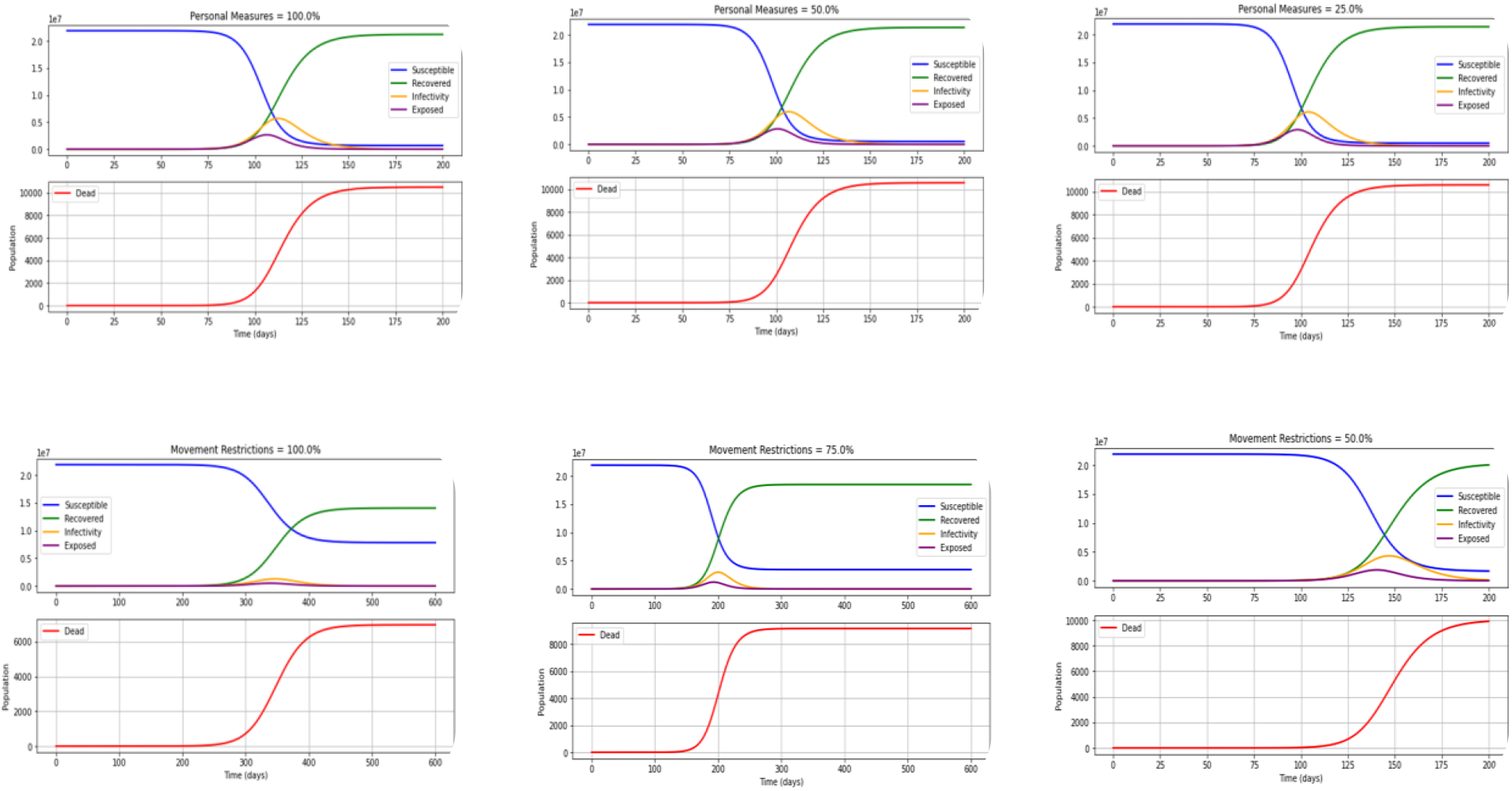

### With implementation of 100%, 75% and 50% of movement restrictions

With the 100%, 75% and 50% of movement restrictions are implemented at R_0_ of 4, the rate of deaths will be increased from 250 days, 150 days, and 110 days with 6,950 deaths in 500 days (with 1.3 million infected individuals), 9,142 deaths in 250 days (with 2.96 million infected individuals) and 9,912 deaths in 200 days (with 4.3 million infected individuals) will be observed. Moreover, the number of days to achieve the peak of the infection curve will be 350 days, 200 days, and 150 days with infected individuals of 0.5 million, 2.5 million and 4 million at the peak of the infection curves, respectively. Refer figure 5 and table 2 for more shows in more details.

### With the implementation of 100%, 50% and 33% of closure of places

With the 100%, 50% and 33% of closure of places are implemented at R_0_ of 4, the rate of deaths will be increased from 110 days, 90 days, and 80 days with 9,912 deaths in 200 days (with 4.3 million infected individuals), 10,417 deaths in 150 days (with 5.4 million infected individuals) and 10,502 deaths in 150 days (with 5.7 million infected individuals) will be observed, respectively. Furthermore, the number of days to achieve the peak of the infection curve will be 150 days, 120 days, and 110 days with an infected individuals of 4 million, 5 million and 6 million at the peak of the infection curves, respectively. Refer figure 5 and table 2 for more shows in more details.

## Discussion

As the COVID-19 pandemic progresses, countries are increasingly implementing a broad range of response activities (Imperial College London, 2020). The present study revealed that it will be necessary to layer multiple interventions, regardless of whether suppression or mitigation is the overarching policy goal. The choice of interventions ultimately depends on the relative feasibility of the implementation of the different strategies and their effectiveness. The compartment models were invented during the late 1920s, which are the most used models in epidemiology. Moreover, different approaches using agent-based simulations are still based on compartment models (Kopp, 2020). The SEIR model is very frequently used to explain the COVID-19 pandemic, which is basic and a reasonably good fit for the disease (He et al., 2020). Results of our paper after simulating various scenarios indicate that disregarding social distancing and hygiene measures can have devastating effects on the Sri Lankan population. However, that model also shows that quarantine of contacts and isolation of cases can help halt the spread of novel coronavirus (Avery et al., 2020). The accuracy of the predictions of the epidemiological models depends critically on the quality of the data feed into the model. If the data quality is good, the model can precisely describe the situations. A fitting example would be when accurately estimating the case fatality rate, which requires all cases of the disease and the number of dead (Adiga et al., 2020). However, during the COVID-19 pandemic, the number of deaths has often been highly inaccurate for many reasons, and the number of infected has also been incorrect. There can be undiagnosed cases during that period because of limited testing, which lead to inaccurate reporting (Adiga et al., 2020; Ferguson et al., n.d.). Furthermore, one of the significant limitations of the model is that it does not include the natural death and birth rates assuming those are constant (Adiga et al., 2020; Mwalili et al., 2020). In addition, during the COVID-19 pandemic, there are broad variations in estimations of Case Fatality Rate (CFR) that may be misleading. Countries may be more or less likely to detect and report all COVID-19 deaths. Furthermore, they may be using different case definitions and testing strategies or counting cases differently. Variations in CFR also may be explained in part by the way time lags are handled. Differing quality of care or interventions being introduced at different stages of the illness also may play a role. Finally, the profile of patients may vary between countries (World Health Organization, 2020b).

The proposed model uses the predictors as given in the parameter table under the methodology section. The model was internally validated using the parameters available in the previous studies in the underpinning literature. As with any modelling approach, our findings relate to the assumptions and inputs of the model which lead to a major limitation. The assumptions with the greatest potential effect on our findings are the structural assumptions of a compartmental epidemiological model (Adiga et al., 2020). Furthermore, the predictive capability of the tool is highly dependent on several preliminary data for parameter estimation. This dependence may lead to data misinterpretation, especially considering the SIR model. Notably, an essential parameter in epidemic modelling is the ‘basic reproduction ratio (R_0_)’. The size of the R_0_ can be varied since it is determined by averaging many cases. Moreover, R_0_ depends on the contagiousness of the pathogen and the number of contacts of an infected person^18^. Furthermore, a study has found an R_0_ of 2.53, implying that the pandemic will persist in the human population in the absence of strong control measures^5^. The parameters, which are locally informed, form the basis of predicting and forecasting exercises accounting for different scenarios and impacts of COVID-19 transmission.

The internal, and external validation of the model is vital for the robust prediction of the ODEs in the model. Thus, the models were applied in the series of equations to get the equilibrium in the SEIRD model. Thereafter, the simulation of the validated model was performed to obtain the policy scenarios of the proposed model. Initially, the model comprised of one exposed individual, and the rest of the population was considered as a susceptible population (Jamrozik & Selgelid, 2020; Nadler et al., 2020; Walmsley et al., 2020). Therefore, the predictors were handled with care in the model to avoid overestimation or underestimation. In addition, the infection fatality ratio (IFR) of COVID-19 acts as a simple factor in the mortality effects of preventive strategies and does not alter the presented relative conclusions. There are limited serological studies to calculate IFR accurately during outbreaks. In such situations, estimates need to be made with routinely available surveillance data, which generally consist of time series of cases and deaths reported in aggregate(World Health Organization, 2020b). When the available data was considered, the situation was almost like a similar study done in China (Verity et al., 2020). The high mortality rate in the COVID-19 pandemic requires that our model have a designated compartment for deaths. The fatality compartment is the only compartment of the model with no further interaction with the rest of the epidemic system. Beta, the proportion between the rate of infection and the rate of spread (R_0_) was predicted when R_0_ equals 4. It is found that the peak of deaths in Sri Lanka may arrive after five months (150 days) following exposure with a maximum number of deaths around 10,617 if there are no preventive measures during the current wave. With high weight to the personal protective measures, the occurrence of deaths will be reduced by 68% and 71% reduction of the infected cases than without having any measures. With high weight to movement restrictions, 35.7% of deaths and 83% of the infected population will be reduced without having any measures. If we give equal consideration for personal protection, movement restriction and closure, only 10.6% of deaths and 36.5% infected population will be prevented without having any measures.

## Conclusion and Recommendation

In the present work, a computational model for predicting the spread of COVID-19 by the dynamic SIERD model has been proposed. The dynamic model assumes a time-dependent death fraction. Various epidemiological parameters such as time of peak arrival, number of active cases and number of deaths during peak are evaluated for all cases and predictions were made against different preventive measures. The key conclusion that we emphasized from this study is the importance of strict adherence, maintenance, and monitoring of the self-preventive measures properly to minimize the death toll from COVID-19. Policymakers need to streamline the resources that are essential for the smooth functioning of this strategy. Polices can be guided by these results which need to be implemented to lower the total population infected, and deaths which will lead to flattening of the curves.

## Data Availability

Publicly available data was taken from the dash board of HPB, Ministry of Health

## Declaration of competing interest

There is no conflict of interest.

## Ethics Approval

The research design and methodology used only anonymized data sets. This entails that no ethical approval is required.

## Funding

This research did not receive any funding from funding agencies.

## Acknowledgement

We would like to acknowledge Health Promotion Bureau,, Ministry of Health and Government of Sri Lanka.

## Author Contribution

Conceptualization and methodology; RMNUR, MSDW, SPJ, IG, PCW; Software; SPJ; Formal analysis; SPJ and RMNUR; Original draft preparation; RMNUR; Writing; RMNUR, BMWIG, WMPCW; Review, suggestions & editing; MSDW, RMNUR, SPJ, TKT, YA, SB.

## Notes

### Competing Interest Statement

The authors have declared no competing interest.

### Clinical Trial

Not applicable

### Funding Statement

We did not receive any funding from funding agencies for this study.

### Summary of Updates

1. Author list and affiliations updated 2. Introduction 3. Discussion 4. References

